# Association of Smoking Behavior, Intensity, and Time Since Cessation with Epigenetic Aging Biomarkers: Results from NHANES 1999-2002

**DOI:** 10.64898/2025.12.18.25342556

**Authors:** Javier Perez-Garcia, Dennis Khodasevich, Anne K. Bozack, Mary B. Rice, Jamaji C. Nwanaji-Enwerem, Nicole Gladish, Belinda L. Needham, David H. Rehkopf, Andres Cardenas

**Author notes:** Corresponding authors: Javier Perez-Garcia, PhD. and Andres Cardenas, PhD. Department of Epidemiology and Population Health, Stanford University, Stanford, CA, USA.

## Abstract

**Background:** Smoking is a major preventable risk factor for all-cause mortality and disability worldwide. It leads to age-related diseases, but the effects and reversibility of smoking behaviors on different epigenetic clocks are not fully explored.

**Objective:** To characterize the association of epigenetic age acceleration in whole blood with active and secondhand smoking (SHS), smoking intensity, and time since cessation among U.S. adults.

**Methods:** This is a cross-sectional study in adults from the NHANES 1999-2002 survey cycles, a population-based survey representative of the U.S. adult civilian non-institutionalized population. We analyzed 2,320 adults aged ≥50 years, including non-Hispanic White, non-Hispanic Black, Mexican American, and other populations. Those without available self-reported smoking status data were excluded. Smoking exposure was analyzed in terms of self-reported smoking status (current, former, never), intensity (packs in the last month), years since smoking cessation, and SHS (serum cotinine levels: 0.05-10 ng/ml). Epigenetic age was estimated using 12 DNA methylation age biomarkers. Survey-weighted linear models were used to estimate the association of smoking exposure with epigenetic age acceleration while adjusting for confounders and multiple comparisons.

**Results:** We analyzed 1,043 never, 903 former, and 374 current smokers (mean age: 65.1±9.3 years, female: 49.1%). GrimAge2 was 9.1 years (95% CI: 8.0, 10.2) and 2.8 years (95% CI: 2.3, 3.3) higher in current and former smokers, respectively, than in never smokers. Smokers showed an increased pace of aging, with current smokers aging 0.15 (95% CI: 0.13, 0.17) and former smokers 0.04 (95% CI: 0.03, 0.05) additional years per chronological year, and shorter methylation-predicted telomere length (current: -132±19 bp; former: -30±15 bp). Each cigarette pack smoked in the past month was associated with increases of 0.1 years in GrimAge2 and PhenoAge, and 0.01 aged months/year in aging pace. Among former smokers, each year since smoking cessation was associated with a deceleration of -0.14 (GrimAge2) and -0.06 (PhenoAge) years, and -0.03 aged months/year in aging pace. Cotinine analyses supported dose-dependent associations of epigenetic aging with smoking and suggested a 0.8-year increase in GrimAge2 with SHS exposure.

**Conclusions:** Smoking was associated in a dose-dependent manner with accelerated epigenetic aging in former and current smokers. However, epigenetic age acceleration declines with time since smoking cessation among former smokers.

## INTRODUCTION

Cigarette-smoking is a major preventable risk factor for all-cause mortality and disability, mostly related to cancer, cardiovascular, and respiratory causes (1). In 2022, 49.2 million U.S. adults reported current tobacco use (2), with cigarette smoking causing almost 530,000 annual deaths (3). Smoking carries a substantial burden for healthcare systems and society, with $225 billion/year in smoking-attributable health care expenditures in the U.S. (4). Targeted cessation interventions are relevant since certain subgroups carry a substantial burden, such as patients with chronic lower respiratory diseases, where smoking is responsible for $18.9 billion of healthcare costs (5).

Quitting smoking is associated with reduced risk of tobacco-related diseases and premature death, improved quality of life, enhanced immune response, and an estimated gain of 10 years of life (1,6,7). Among cigarette smokers, health benefits from smoking cessation can be realized regardless of the age at which cessation occurs, as reductions in mortality risk are observed even in individuals who discontinue smoking after age 70 (1,8). Despite the evident declines in mortality and disability following smoking cessation and substantial government efforts to reduce smoking (1,9), predictions suggest that smoking will remain a major health concern for at least the next 10 years (9).

There is robust evidence that smoking induces changes in DNA methylation (DNAm), which could contribute to smoking-related disease mechanisms (10). DNAm is an epigenetic process that regulates gene expression without modifying the DNA sequence, with potential impact in disease pathogenesis (11). Smoking alters numerous DNAm loci across the genome (e.g., *AHRR*) implicated in pathologies like asthma, malignancies, neurological, and developmental diseases (10–17). Although tobacco exposure can induce persistent DNAm markers detectable for up to 30 years after smoking cessation, or even persist from *in utero* exposure into adulthood, there is evidence that some markers rapidly attenuate after smoking cessation (10,12–15).

Selected DNAm biomarkers, known as epigenetic clocks, are highly accurate predictors of biological aging. Increased deviation of epigenetic age over chronological age (i.e., epigenetic age acceleration) reflects age-related diseases, such as chronic respiratory diseases (18–21) and cardiometabolic and cardiovascular diseases (22,23), as well as prenatal exposures and pregnancy complications (24,25). Smoking may accelerate epigenetic aging, potentially representing a mechanism underlying the pathogenic effect of smoking and positioning epigenetic clocks as potential biomarkers of its health impact (11,16,17). Nevertheless, the effects of smoking on epigenetic aging are not fully elucidated. There are multiple epigenetic clocks capturing distinct biological processes that have not been evaluated in the context of active and passive smoking, with limited information regarding whether smoking intensity and cessation influence epigenetic aging biomarkers.

Therefore, we aimed to conduct a comprehensive characterization of the association of epigenetic age acceleration in whole blood with active and secondhand smoking (SHS), intensity, and time since cessation among U.S. adults.

## METHODS

### Study population

We analyzed public data from the National Health and Nutrition Examination Survey (NHANES), a biannual program aimed at collecting demographic, medical, behavioral, and nutritional data representative of the U.S. adult civilian noninstitutionalized population (26). DNAm data were generated in a subset of 2,532 adults aged ≥50 years old surveyed in 1999-2000 or 2001-2002 cycles who had whole blood samples for DNA purification. All NHANES participants provided written informed consent, and study protocols were approved by the NCHS Research Ethics Review Board.

### DNA methylation data and aging predictors estimation

A full description of sample collection, laboratory methods, and bioinformatics procedures to compute and analyze epigenetic clocks is described in the supplementary material and NHANES website (27). Briefly, genome-wide DNAm was measured in whole blood using the Infinium MethylationEPIC beadchip (Illumina, San Diego, USA). After quality control, DNAm epigenetic biomarkers were calculated, including chronological age clocks (HorvathAge, HannumAge, SkinBloodAge, LinAge, WeidnerAge, VidalBraloAge, and ZhangAge), lifespan and health span clocks (PhenoAge and GrimAge2), pace of aging (DunedinPoAm), DNAm-estimated telomere length (DNAmTL), and relative estimate of the number of stem cell divisions per cell (epiTOC). Complementarily, telomere length was directly measured in whole blood samples through a quantitative polymerase chain reaction (qPCR) (28).

### Smoking behavior and exposure

Smoking exposure was assessed through self-reported standardized questionnaires administered by trained interviewers. Never, former, and current smoking were defined based on 1) having smoked at least 100 cigarettes in their lifetime and 2) reporting currently active smoking. Former smokers provided information about how long it had been since smoking cessation. Smoking intensity among current smokers was defined based on cigarette packs in the last month. SHS exposure was defined among non-current smokers as those with 0.05-10 ng/ml of serum cotinine levels (29).

### Statistical analyses

We excluded participants aged ≥85 years (n=130) (ages ≥85 were top-coded at 85 years), those with sex discordances between reported and epigenetic-predicted sex (n=56), participants without data about self-reported smoking (n=6), and never cigarette smokers who ever smoked pipe or cigar (n=20), leaving 2,320 participants for subsequent analyses. We examined the association between self-reported smoking status, smoking intensity, time since smoking cessation, and SHS with epigenetic clocks through survey-weighted linear regression models adjusted for age, age^2^, gender, race-ethnicity, body mass index, poverty-to-income ratio, education, and survey cycle (26). Additionally, for comparison with never smokers, we stratified former smokers according to time since smoking cessation (from ≤5 to >30 years) and current smokers according to smoking intensity (from ≤10 to >30 packs/month). We also evaluated qPCR-measured telomere lengths with smoking exposures. Multiple comparisons were adjusted using a false discovery rate (FDR)<5% considering the number of epigenetic clocks evaluated. All analyses were conducted in R (v 4.4.1).

### Sensitivity analyses

We conducted sensitivity analyses for all epigenetic clocks associations by further adjusting the models for methylation-derived blood cell counts (CD8^+^ T cells, CD4^+^ T cells, NK cells, B cells, monocytes, and neutrophils). Furthermore, we used cotinine levels as a measurable biomarker to validate the association of epigenetic aging with self-reported smoking status and intensity. Finally, since DNAm-predicted pack years is a component of GrimAge2, we inspected whether the associations of this clock may be driven by other mortality predictors.

## RESULTS

### Study population

We analyzed 2,320 adults from NHANES 1999-2002, whose demographic characteristics are summarized in **Table 1**. Subjects had a mean age (standard deviation, SD) of 65.1±9.3 years, 49.1% were female, and had a mean BMI of 28.8±5.8 kg/m^2^. They self-identified as Non-Hispanic White (39.1%), Mexican American (29.1%), Black (21.9%), other Hispanic (6.5%), or other/multi-racial (3.4%). Participants were self-reported as never smokers (45.0%), former smokers (38.9%), and current smokers (16.1%). The mean time since smoking cessation was 19.5±13.5 years among former smokers. Based on serum cotinine levels, 34.7% of never smokers were predicted to be exposed to SHS. Current smokers reported smoking 21.9±16.9 cigarette packs in the last month.

**Table 1.**
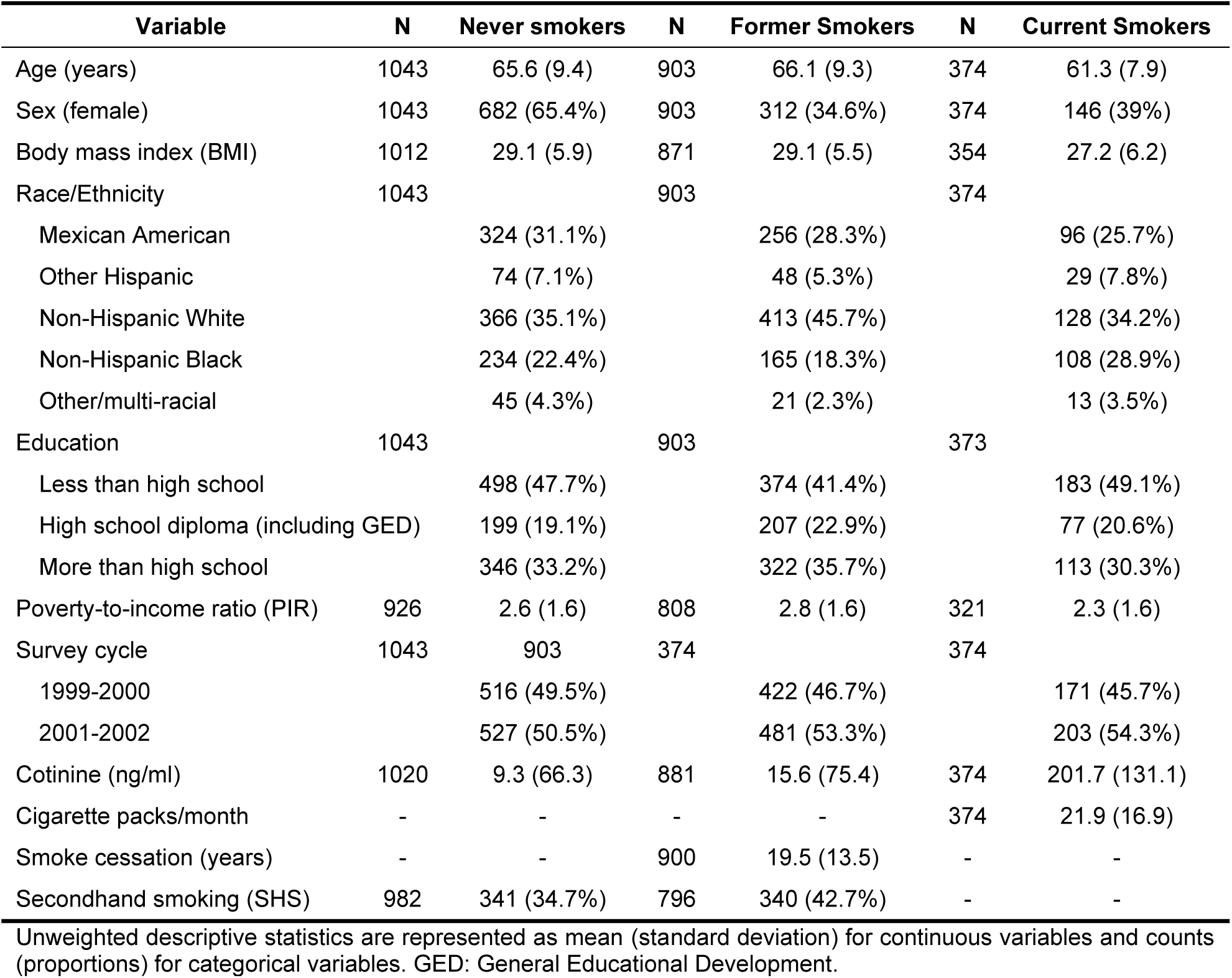
Unweighted Sample Characteristics of NHANES 1999-2002 participants included in the study stratified by smoking status.

Most of the epigenetic clocks showed high correlation with chronological age in the analyzed participants, except for DunedinPoAm, which, as expected, was uncorrelated with age, and epiTOC, which showed a modest correlation (**Figure S1**).

### Self-reported smoking status is associated with epigenetic aging

Current and former smokers showed accelerated epigenetic aging across several biomarkers compared to never smokers, with a notable effect among current smokers (**Figure 1**, **Table S1**). GrimAge2 was 9.1 years higher (95% Confidence Interval [CI]: 8.0, 10.2) and PhenoAge 2.4 years higher (95% CI: 1.0, 3.8) in current smokers compared to never smokers. Current smokers also had -132 bp (95% CI: -169, -96) shorter DNAmTL and a faster pace of aging of 0.15 (95% CI: 0.13, 0.17) additional years of physiological decline per chronological year compared to never smokers. Similarly, GrimAge2 was 2.8 years higher (95% CI: 2.3, 3.3) and the pace of aging was 0.04 years of physiological decline/calendar year higher (95% CI: 0.03, 0.05) in former smokers compared to never smokers. Only WeidnerAge showed a negative association (-2.2 years; 95% CI: -3.6, - 0.8) with current smoking status compared to never smokers.

**Figure 1.**
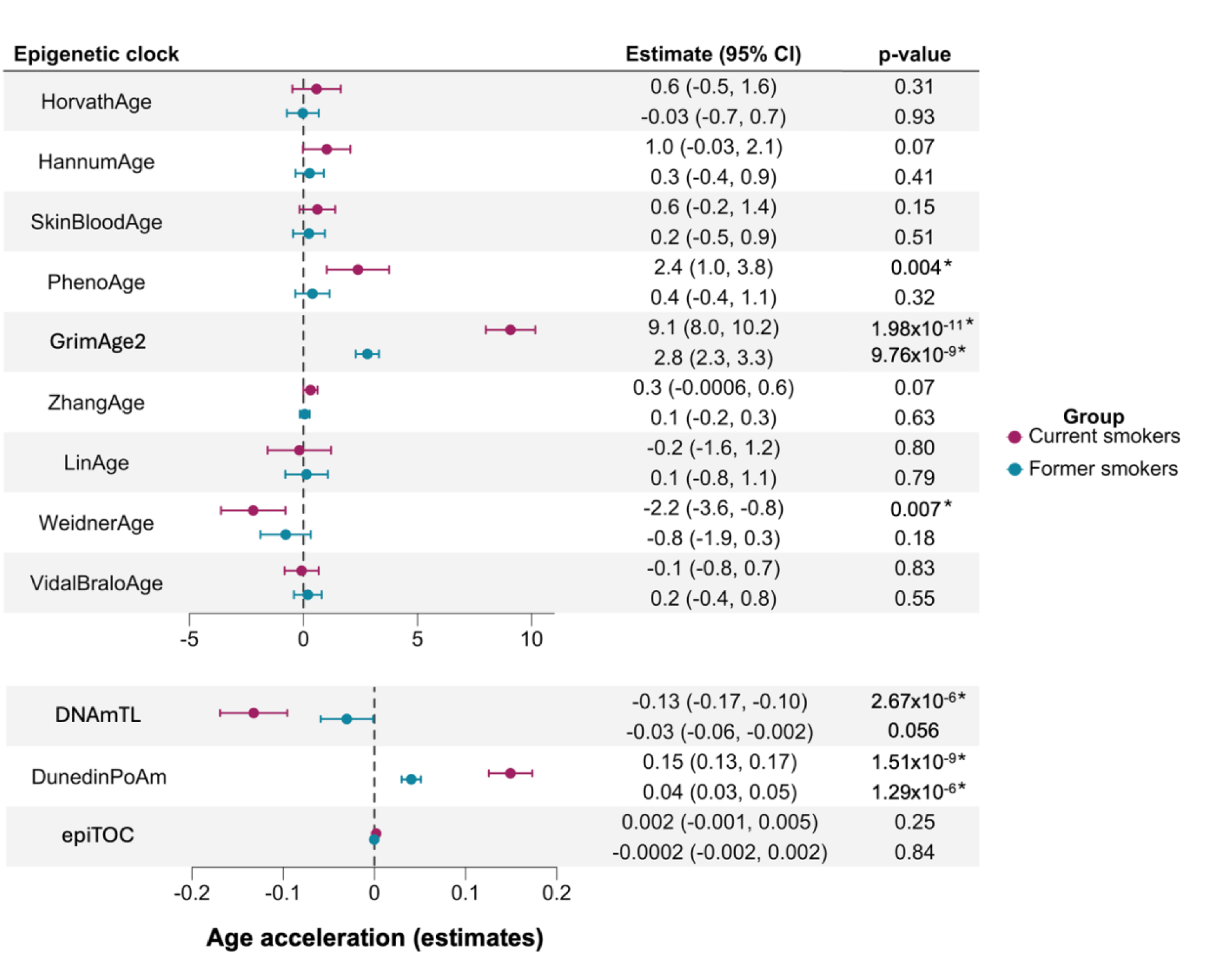
Forest plot of the association of epigenetic clocks with self-reported smoking status, for current (pink) and former (blue) smokers relative to never smokers. Effect estimates and 95% confidence intervals are represented on the x-axis. Effect estimates for HorvathAge, HannumAge, SkinBloodAge, PhenoAge, GrimAge2, ZhangAge, LinAge, WeidnerAge, and VidalBraloAge are in years; for DNAmTL are in kilobases; for DunedinPoAm are in unit differences of the pace of aging (a score of 1 represents an average aging pace); and for epiTOC are in average beta values in CpGs that represent a relative estimate of the number of stem cell divisions per stem cell. Positive estimates indicate accelerated epigenetic aging, except for DNAmTL, since shorter telomere lengths indicate accelerated aging. The vertical dashed line at zero represents no association. *False discovery rate<0.05.

### Smoking intensity is associated with accelerated epigenetic aging

Among current smokers, each additional cigarette pack smoked in the last month was associated with 0.1 years (95% CI: 0.04, 0.2) higher GrimAge2 (95% CI: 0.04, 0.2), higher pace of aging of 0.001 (95% CI: 4×10^-4^, 0.002) additional years of physiological decline/calendar year, and -2.4 bp (95% CI: -3.8, -1.0) shorter DNAmTL (**Figure 2A, Table S2**). When current smokers were stratified by smoking intensity, epigenetic age acceleration increased compared with the number of cigarette packs smoked in the last month, compared to never smokers (**Figure 2B, Table S3**).

**Figure 2.**
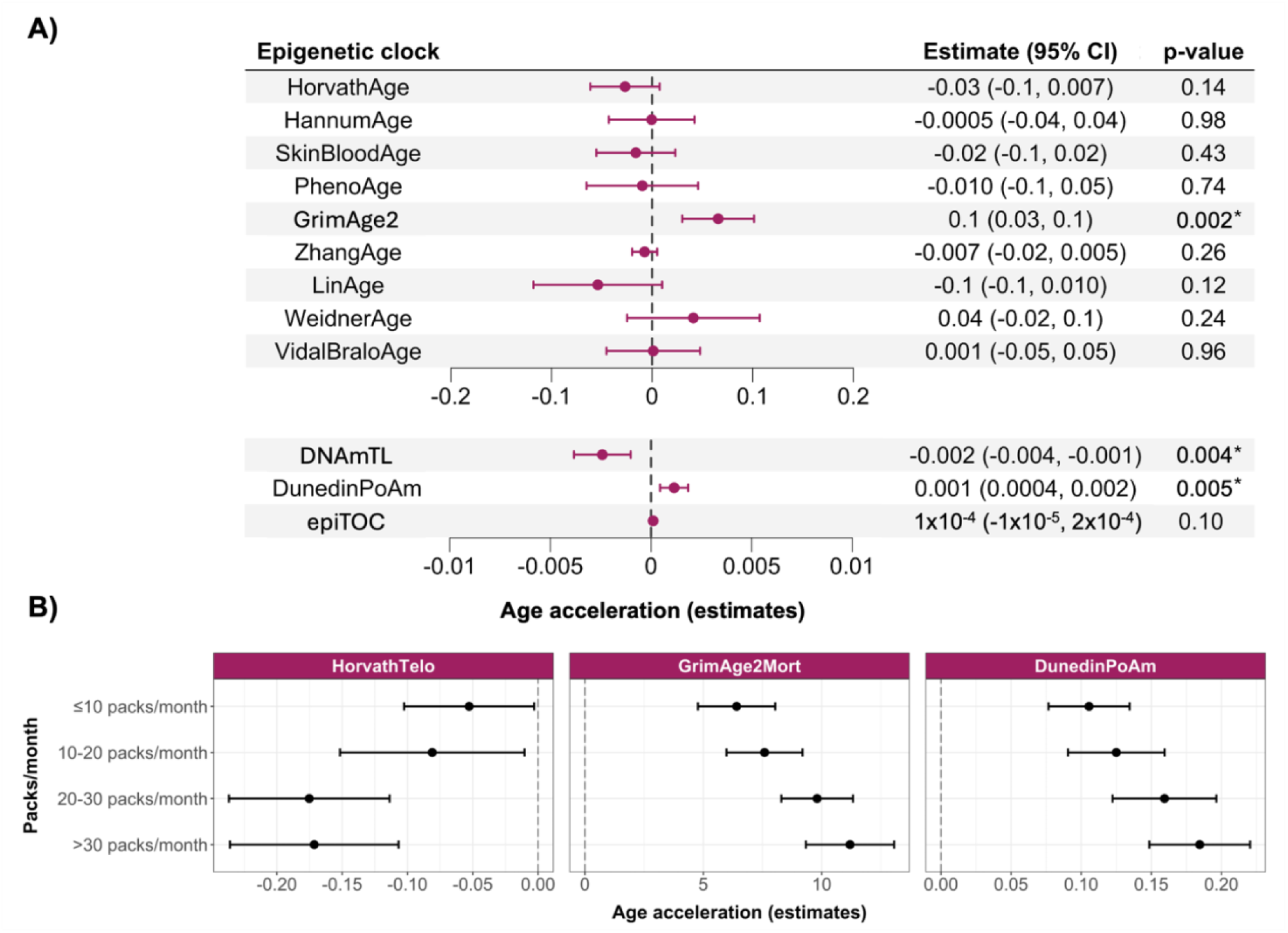
Forest plots of **A)** the association of epigenetic clocks with smoking intensity (packs in the last month) among current smokers and **B)** the comparison of epigenetic clocks between never and current smokers stratified by smoking intensity. Effect estimates and 95% confidence intervals are represented on the x-axis. Effect estimates for HorvathAge, HannumAge, SkinBloodAge, PhenoAge, GrimAge2, ZhangAge, LinAge, WeidnerAge, and VidalBraloAge are in years; for DNAmTL are in kilobases; for DunedinPoAm are in unit differences of the pace of aging (a score of 1 represents an average aging pace); and for epiTOC are in average beta values in CpGs that represent a relative estimate of the number of stem cell divisions per stem cell. Positive estimates indicate accelerated epigenetic aging, except for DNAmTL, since shorter telomere lengths indicate accelerated aging. The vertical dashed line at zero represents no association. *False discovery rate<0.05.

Heavy smokers’ (>30 packs/month) GrimAge2 was 11.2 years (95%CI: 9.3, 13.1) higher compared to never smokers, while light smokers’ (≤10 packs/month) GrimAge2 was 6.4 years (95%CI: 4.8, 8.0) higher. Additionally, DNAmTL was -171 bp shorter (95% CI: -236, -107), and the pace of aging was 0.18 (95% CI: 0.15, 0.22) aged physiological years per calendar year higher in heavy smokers compared to never smokers. Similarly, light smokers exhibited shorter DNAmTL of -52.8 bp (95% CI: -102.6, -3.0) and a 0.11-increase (95% CI: 0.08, 0.13) in their pace of aging.

### Time since smoking cessation is associated with lower epigenetic aging

Among former smokers, each additional year since smoking cessation was associated with a lower age acceleration of -0.14 years in GrimAge2 (95% CI: -0.17, - 0.11) and -0.06 years in PhenoAge (95% CI: -0.09, -0.04), and a slower pace of aging of -0.002 additional years of physiological decline per calendar year (95% CI: -0.003, -0.002) (**Figure 3A**, **Table S4**). Each additional year since smoking cessation was nominally associated with a 2.5-bp increase in DNAmTL (95% CI: 0.6, 4.5, *p*=0.02, FDR=0.07).

**Figure 3.**
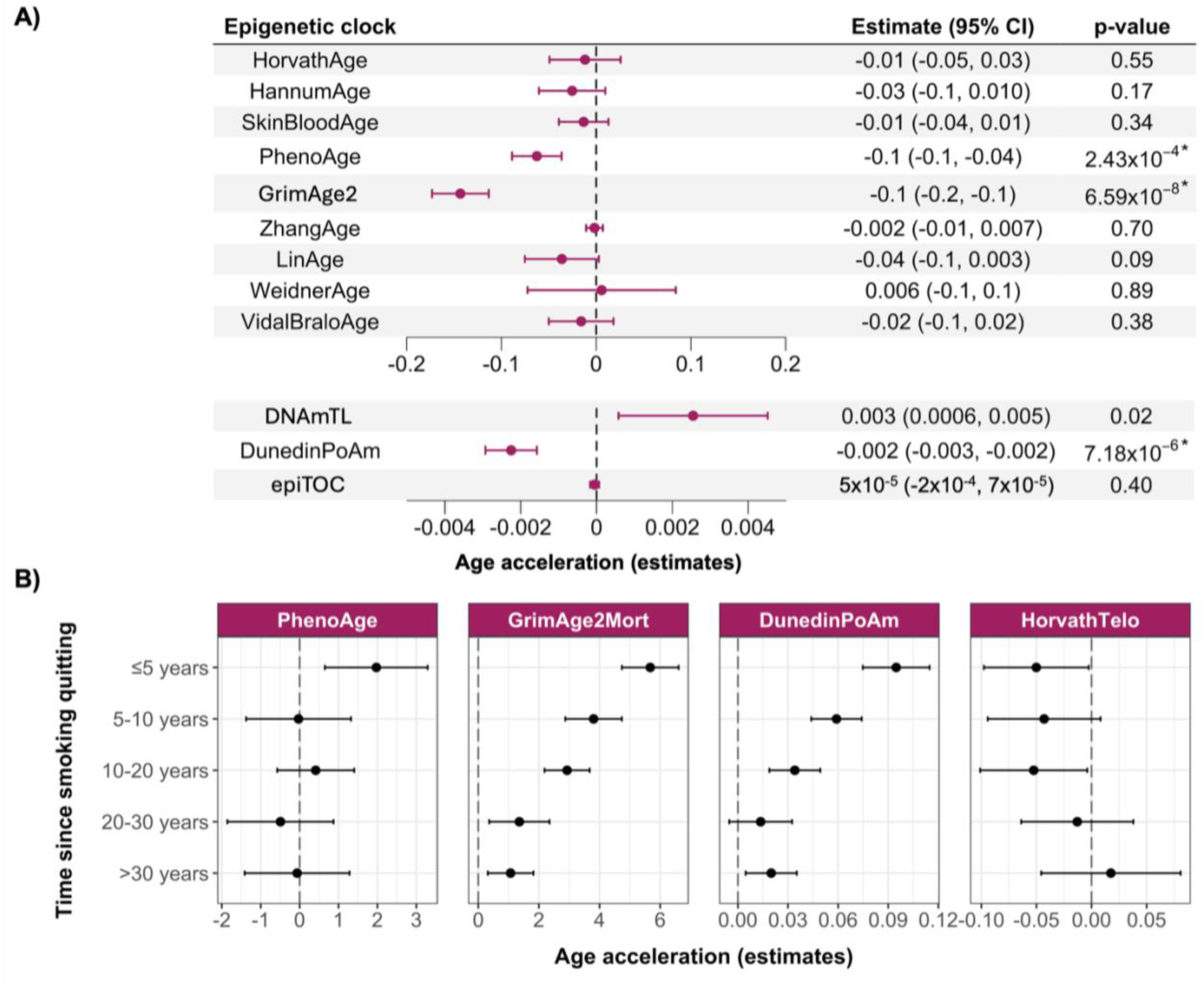
Forest plots of **A)** the association of epigenetic clocks with time since smoking cessation in years among former smokers and **B)** the comparison of epigenetic clocks between never smokers and former smokers stratified by time since smoking cessation. Effect estimates and 95% confidence intervals are represented on the x-axis. Effect estimates for HorvathAge, HannumAge, SkinBloodAge, PhenoAge, GrimAge2, ZhangAge, LinAge, WeidnerAge, and VidalBraloAge are in years; for DNAmTL are in kilobases; for DunedinPoAm are in unit differences of the pace of aging (a score of 1 represents an average aging pace); and for epiTOC are in average beta values in CpGs that represent a relative estimate of the number of stem cell divisions per stem cell. Positive estimates indicate accelerated epigenetic aging, except for DNAmTL, since shorter telomere lengths indicate accelerated aging. The vertical dashed line at zero represents no association. *False discovery rate<0.05.

Moreover, associations of epigenetic aging among former compared to never smokers were mitigated with increasing time since smoking cessation (**Figure 3B, Table S5**). While GrimAge2 was 5.7 years higher (95%CI: 4.7, 6.6) and PhenoAge 2.0 years higher (95%CI: 0.6, 3.3) in recent quitters (≤5 years) compared to never smokers, long-term quitters (>30 years) had GrimAge2 only 1.1 years higher (95% CI: 0.3, 1.8) and PhenoAge was indistinguishable from never smokers (*p*=0.93). Recent quitters had a faster pace of aging than never smokers (0.09 additional years of physiological decline per calendar year; 95% CI: 0.07, 0.11) and a suggestively shorter DNAmTL (-50.1 bp; 95%CI: -97.7, -2.5; *p*=0.06). Conversely, long-term quitters showed a smaller increase in the pace of aging (0.02 units; 95% CI: 0.005, 0.04) and no differences in DNAmTL compared with never smokers (*p*=0.59).

### Association between secondhand smoking and epigenetic aging

No epigenetic clock was associated with SHS exposure among never smokers (*p*>0.05; **Table S6**). However, when analyzing never and former smokers who quit smoking >1 month ago, we found that those exposed to SHS had an 0.8-year higher GrimAge2 relative to non-exposed (95% CI: 0.3, 1.2; *p*=0.009; FDR=0.11).

### Smoking is not associated with qPCR-measured telomere length

No significant associations were observed between qPCR-measured leukocyte telomere lengths and smoking status, intensity, or time since smoking cessation (*p*>0.05; **Table S7**).

### Epigenetic aging is associated with cotinine levels as a proxy of smoking behavior

Independent of self-reported smoking status, higher serum cotinine levels were significantly associated with higher PhenoAge, DunedinPoAm, and GrimAge2 and with shorter DNAmTL (FDR<0.05; **Figure 4**, **Table S8**). Additionally, serum cotinine was suggestively associated with higher HannumAge and SkinBloodAge (acceleration of 0.3-1.0 years per SD increment in cotinine levels; *p*=0.03; FDR=0.05). When stratified by self-reported smoking status, higher cotinine levels among current smokers (as a biomarker of recent smoking intensity) were associated with accelerated PhenoAge, DunedinPoAm, and GrimAge2 and with shorter DNAmTL (*p*<0.05; **Table S9**).

**Figure 4.**
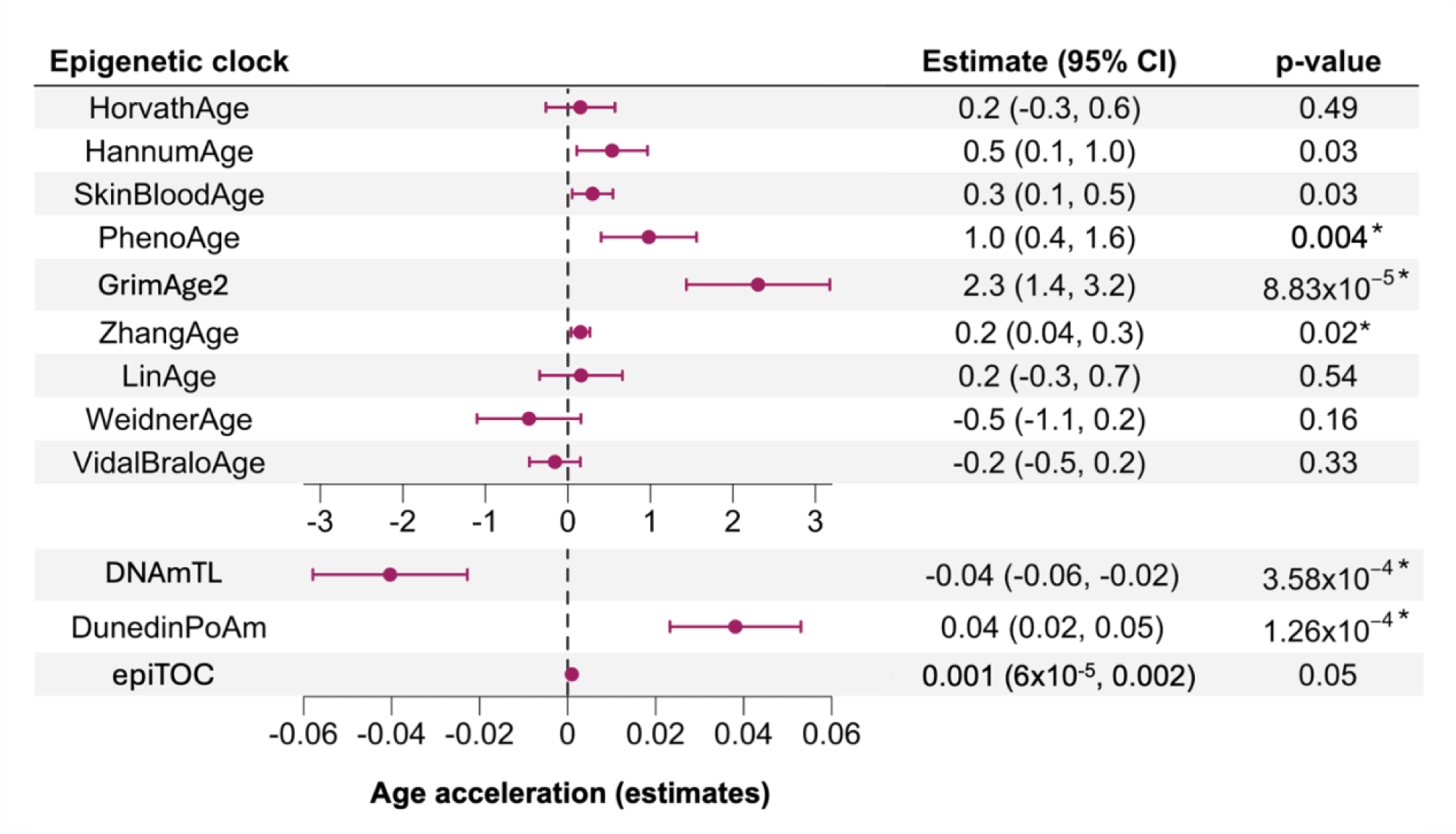
Forest plot of the association of epigenetic clocks with serum cotinine levels (ng/ml). Effect estimates and 95% confidence intervals (per one standard deviation increase) are represented on the x-axis. Effect estimates for HorvathAge, HannumAge, SkinBloodAge, PhenoAge, GrimAge2, ZhangAge, LinAge, WeidnerAge, and VidalBraloAge are in years; for DNAmTL are in kilobases; for DunedinPoAm are in unit differences of the pace of aging (a score of 1 represents an average aging pace); and for epiTOC are in average beta values in CpGs that represent a relative estimate of the number of stem cell divisions per stem cell. Positive estimates indicate accelerated epigenetic aging, except for DNAmTL, since shorter telomere lengths indicate accelerated aging. The vertical dashed line at zero represents no association. *False discovery rate<0.05.

### Sensitivity analyses

The GrimAge2 components GDF15, CystatinC, ADM, PAI-1, Leptin, CRP, logHA1C, and TIMP-1 were significantly associated with smoking status, time since cessation, intensity, and/or cotinine levels (*p*<0.05, **Table S10**). All associations of accelerated epigenetic aging with smoking exposures remained significant after further adjustment by cell type composition (*p*<0.05; **Table S1-S6, S8-S10**).

## DISCUSSION

To the best of our knowledge, this is the first study to comprehensively investigate associations between active and SHS smoking, intensity, and time since cessation with 12 validated epigenetic aging biomarkers in a nationally representative sample of U.S. civilian noninstitutionalized adults. We found that smokers showed accelerated epigenetic aging across several biomarkers compared to never smokers, with a more pronounced effect among current than former smokers. Smoking tobacco was associated with epigenetic aging acceleration in a dose-dependent manner in clocks reflecting morbidity, mortality, pace of aging, and telomere shortening. Notably, epigenetic age acceleration was attenuated with longer time since quitting among former smokers, suggesting its reversibility. Analyses of serum cotinine levels confirmed associations between epigenetic aging and smoking status and intensity, while no substantial effect was observed for SHS exposure.

We reported accelerated aging among smokers compared to never smokers, mainly in second- and third-generation epigenetic clocks, reflecting morbidity/mortality and aging rates, respectively. Among current smokers, accelerated aging was reflected in an increase of up to 2-9 years in PhenoAge and GrimAge2, and in a faster pace of aging corresponding to 0.15 additional years of physiological decline per calendar year. These results are consistent with the existing literature, where smoking has been predominantly associated with mortality, morbidity, and physiological decline clocks, while chronological clocks are less responsive to smoking (11,16,17,30). This is expected since first-generation clocks were not built to capture environmental variation, such as smoking, to minimize variations among same-age subjects (16,31). Only WeidnerAge showed an inverse association with smoking, similar to that described in a previous study (32). We hypothesize that this association might be an artifact related to this clock rather than smoking having a protective effect. It was trained for forensic analyses using bisulfite pyrosequencing in relatively small datasets unbalanced for sex and cell types (33), and exhibited the lowest correlation with chronological age in our dataset.

Nevertheless, when analyzing serum cotinine as a biomarker of recent smoking exposure, we reported associations with two chronological epigenetic clocks, HannumAge and SkinBloodAge. In large epidemiological studies, self-reporting bias may underestimate the smoking patterns in the sample, suppressing the associations with epigenetic age acceleration (34). Cotinine serves as a complementary biomarker that objectively provides information about active smoking but also reflects recent exposure intensity, SHS exposure, and variability in nicotine metabolism (35,36). Our results suggest that actual smoking exposure, beyond self-reporting behavior, may influence biological aging in a way that could be captured by predictive clocks of chronological age, contrary to previous assumptions. Combining self-reported smoking, which reflects cumulative long-term exposure, with cotinine levels, a complementary short-term biomarker, would provide more informative markers of smoking-related biological aging.

Furthermore, we observed that active smoking was associated with accelerated epigenetic aging in an intensity-dependent manner. The degree of accelerated epigenetic aging increased proportionally with the number of cigarettes smoked in the last month and serum cotinine levels among current smokers. We estimated that each cigarette smoked in the past month would contribute to an aging acceleration of 1.3 days in GrimAge2. While light smokers (≤10 packs/month) were 6 years older in GrimAge2 and had an accelerated aging pace of 1.3 months/calendar year compared to never smokers, heavy smokers (>30 packs/month) were 10 years older and aged 2.2 months per year faster. Our findings agree with two population-based studies and two in adult women, where pack-years or cigarettes/day were associated with mortality (GrimAge2) and/or pace of aging clocks (37–40).

However, we observed greater epigenetic age acceleration in current than former smokers, supporting a potential reversibility after cessation as previously suggested (41–44). We found that each year of smoking cessation was associated with a deceleration of 1-2 months in mortality clocks, accompanied by a slower pace of aging and lower DNAmTL reductions. While recent quitters were up to 5 years older in mortality clocks, long-term quitters showed minimal or no differences in epigenetic age. Notably, we observed clock-specific recovery patterns after cessation. While a clinical-phenotype-based clock (PhenoAge) quickly normalized within ∼5-10 years after cessation, DNAmTL, as a reflection of cell replicative history, showed an intermediate recovery (∼10-30 years). However, mortality and physiological decline clocks (GrimAge2 and DunedinPoAm) do not converge to never smokers until approximately >20-30 years. The reversibility of smoking DNAm damage is tissue-dependent (45), with our findings adding that it is also biomarker-dependent, with different clocks providing complementary information on smoking-related biological aging and recovery. These promising results could further strengthen anti-smoking policies and provide additional motivation for individuals to quit smoking.

Moreover, SHS was suggestively associated with accelerated epigenetic aging among former and never smokers, although no effect was observed when analyzing never smokers alone. This may be attributable to former smokers being more likely to reside in environments where they cohabit with active smokers (46), resulting in more sustained exposure to SHS. Indeed, former smokers had higher serum cotinine levels than never smokers (15.6 *vs.* 9.3 ng/ml, *p*=2.2×10^-8^). There is substantial evidence that SHS can induce DNAm changes and accelerate epigenetic aging, particularly for *in-utero* and childhood exposures (47–51). Our study suggests that DNAm could capture the biological aging related to SHS in adults, similar to other aging biomarkers (52,53).

Finally, most of the evaluated biomarkers and DNAm-based GrimAge2 components showed a negative impact of smoking on biological aging and potential reversibility after cessation, except for β-2 microglobulin (B2M) and qPCR-based telomere lengths. All GrimAge2 components, except B2M, were associated with current smoking and/or cotinine levels, supporting that GrimAge2 associations were not driven only by DNAm-predicted pack-years. This lack of association in our study could be related to variation in B2M plasma levels due to factors such as age or kidney function. Furthermore, adrenomedullin (ADM), growth differentiation factor 15 (GDF15), hemoglobin A1c (HbA1c), plasminogen activation inhibitor 1 (PAI-1), and tissue inhibitor metalloproteinase 1 (TIMP-1) were also associated with smoking intensity and cessation. These contribute to hypertension and heart failure, cognitive dysfunction, age-related mitochondrial dysfunction, anti-apoptosis processes, and lifespan (54), suggesting a potential reversible effect of smoking on these mortality-related traits. Moreover, although no significant association was observed when analyzing qPCR-based telomere length, DNAm-based predictions capture independent biological variation relevant to health outcomes, immune-senescence, and mortality (55,56).

Our study has major strengths. First, we analyzed a large DNAm dataset from a representative sample of the U.S. adult population. Secondly, we analyzed first, second, and third-generation epigenetic clocks reflecting different aspects of epigenetic aging derived from high-quality DNAm data. Third, we employed rigorous statistical methods to address survey-design biases, potential confounders, and multiple comparison issues to minimize false positives.

However, several limitations must be acknowledged. First, although NHANES is a highly valuable source of health data from the U.S. population, it mostly relies on self-reported data, which may introduce some biases in exposure measurements. However, we partially addressed this limitation by analyzing cotinine levels. Second, as NHANES is a cross-sectional study, longitudinal approaches will be needed to confirm that smoking cessation can reverse the effect of smoking on epigenetic age. Third, although DNAm is among the most useful biomarkers of biological age, aging is a complex trait, and its study may be complemented with other omics in different tissues.

## CONCLUSIONS

We found that former and current smoking, assessed by self-report and serum cotinine levels, were associated with accelerated epigenetic aging. We also found evidence to suggest that SHS exposure may accelerate epigenetic mortality clocks. Among current smokers, epigenetic aging proportionately increased with each pack smoked in the past month. However, among former smokers, this association was attenuated with each year since smoking cessation, suggesting a degree of reversibility.

## ABBREVIATIONS

ADM: Adrenomedullin
B2M: β-2 microglobulin
CRP: High-sensitivity C-reactive protein
DNAm: DNAmTL
DNA: methylation
DNA: methylation estimated telomere length
GDF15: Growth differentiation factor 15
logHA1C: Log scale of hemoglobin A1c
NHANES: National Health and Nutrition Examination Survey
PACKYRS: Pack years of smoking
PAI-1: Plasminogen activation inhibitor 1
SHS: Secondhand smoking
TIMP-1: Tissue inhibitor metalloproteinase 1

## AUTHOR CONTRIBUTIONS

JP-G and AC were involved in the conceptualization and design of the study; JP-G, DK, AB, NG, and AC in data curation, formal analyses, and/or data interpretation; SDM and MBR helped in the interpretation of results and manuscript writing; NG, BLN, and DHR generated the data, supervised initial biomarker calculations, and/or secured funding; JP-G leads the visualization and writing of the original draft; and AC was responsible for funding acquisition and project supervision. All authors were involved in the critical revision of the manuscript and have read and agreed to the published version of the manuscript. The authors agree to be accountable for all aspects of the work, ensuring its scientific accuracy and integrity.

## DATA SHARING STATEMENT

Demographic data, health records, and DNA methylation epigenetic clocks estimated in the analyzed participants are publicly available on the NHANES website. The full summary statistic report of all association analyses supporting the main conclusions of this study, including main analyses, sensitivity analyses, as well as the code for data analysis, will be openly available in the Supplementary Material of this manuscript and in the Zenodo repository (DOI:10.5281/zenodo.17977043) after the publication of the manuscript.

## ACKNOWLEDGMENTS

The authors thank all the participants in the NHANES 1999-2000 and 2001-2002 cycles, as well as all researchers and people involved in the study design, data collection, data curation, sample processing, methylation data generation, and open data publication.

