## Supplementary Material for "Association of Smoking Behavior, Intensity, and Time Since Cessation with Epigenetic Aging Biomarkers: Results from NHANES 1999-2002"

This original research article contains the following Supplementary Material. Note that Supplementary Tables are uploaded in an independent file.

**TABLE OF CONTENTS**


| | **Pages** |
|---|---|
| **Supplementary Methods** | 3-7 |
| **Supplementary References** | 8 |
| **Supplementary Figures** | |
| **Figure S1.** Scatter plot and correlation between chronological age and epigenetic clocks. | 9-10 |
| **Supplementary Tables** | |
| **Table S1.** Associations of smoking status with epigenetic clocks. | |
| **Table S2.** Associations of smoking intensity (packs in the past month) with epigenetic clocks. | |
| **Table S3.** Comparison of epigenetic clocks between never and current smokers stratified by smoking intensity. | |
| **Table S4.** Associations of time since smoking cessation with epigenetic clocks. | |
| **Table S5.** Comparison of epigenetic clocks between never and former smokers stratified by time since smoking cessation. | |
| **Table S6.** Associations of secondhand smoking in never smokers with epigenetic clocks. | |
| **Table S7.** Associations of smoking exposures with telomere length. | |
| **Table S8.** Associations of serum cotinine levels with epigenetic clocks. | |
| **Table S9.** Associations of serum cotinine levels with epigenetic clocks only among current smokers. | |
| **Table S10.** Associations of smoking exposures with GrimAge2 components. | |

### SUPPLEMENTARY METHODS

#### Study population

We analyzed public data of participants from the National Health and Nutrition Examination Survey (NHANES) 1999-2000 and 2001-2002 cycles. NHANES is a biannual program coordinated by the National Center for Health Statistics (NCHS) aimed at collecting demographic, medical, behavioral, and nutritional data representative of the U.S. adult civilian noninstitutionalized population (1). Epigenome-wide DNAm data were generated in a subset of 2,532 NHANES participants. The inclusion criteria were adults aged >50 years old surveyed in 1999-2000 or 2001-2002 cycles who had whole blood samples for DNA purification. The sample included a random selection of approximately half of the eligible non-Hispanic White participants and all the eligible non-Hispanic Black, Mexican American, other Hispanic, and other race participants. All NHANES participants provided written informed consent, and study protocols were approved by the NCHS Research Ethics Review Board.

#### DNA methylation data and epigenetic clocks estimation

A full description of sample collection, laboratory methods, and bioinformatics procedures used to process the NHANES DNAm array data and compute epigenetic clocks in blood is described on the NHANES website (2). Briefly, genomic DNA was extracted from whole blood samples and treated with sodium bisulfite using the Zymo EZ DNA Methylation Kit (Zymo Research, Irvine). DNAm in more than 850.000 CpGs across the genome was measured using the Illumina Infinium MethylationEPIC beadchip (Illumina, San Diego, CA, USA).

After pre-processing and quality control of array-derived data, multiple DNAm epigenetic biomarkers were estimated. These included chronological age clocks (HorvathAge, HannumAge, SkinBloodAge, LinAge, WeidnerAge, VidalBraloAge, and ZhangAge), lifespan and health span clocks (PhenoAge and GrimAge2), pace of aging (DunedinPoAm), DNAm telomere length (DNAmTL), and relative estimate of the number of stem cell divisions per stem cell (epiTOC). Multiple GrimAge predictors of time of death were estimated, including adrenomedullin (ADM), β-2 microglobulin (B2M), cystatin C (CystatinC), growth differentiation factor 15 (GDF15), leptin (Leptin), hemoglobin A1c (logHA1C), high-sensitivity C-reactive protein (CRP), plasminogen activation inhibitor (PAI-1), tissue inhibitor metalloproteinase 1 (TIMP-1), and pack years of smoking (PACKYRS). The correlation between chronological and epigenetic age was inspected through the Spearman correlation test.

Additionally, proportions for six blood cell types (neutrophils, monocytes, B-lymphocytes, natural killer cells, $CD4^+$, and $CD8^+$ cells) were estimated using the IDOL probe subset in combination with the FlowSorted.Blood.EPIC reference dataset using the *estimateCellCounts2* function from the *FlowSorted.Blood.EPIC* R package (3).

**Smoking behavior and exposure**

Data about smoking behavior and exposure were recorded through self-reported standardized questionnaires administered by trained interviewers and retrieved from the NHANES website (question identifiers) (1). Smoking status was defined based on self-reported cigarette smoking behavior throughout life (SMQ020), based on the Centers for Disease Control (CDC) classification, and current smoking (SMQ040). Never smoking was defined as not having smoked at least 100 cigarettes in life and not having smoked a pipe (SMQ120) or cigars (SMQ150). Former smoking was defined as having smoked at least 100 cigarettes in life but not reporting current smoking, while current smoking was defined as having smoked at least 100 cigarettes in life and reporting current active smoking. Former smokers provided information about how long it has been since smoking cessation (SMQ050Q, normalized by the time unit of measure reported in SMQ050U). Among current smokers, we estimated the cigarette packs per month as an approximation of smoking intensity based on the self-reported estimation of cigarettes smoked per day (SMD090) and the number of days in the past month that participants smoked a cigarette (SMD080).

Environmental tobacco smoke exposure was also assessed through the measurement of serum cotinine (LBXCOT), a major metabolite of nicotine marker for both active and passive smoking. Serum cotinine (ng/ml) was measured by an isotope dilution-high performance liquid chromatography and atmospheric pressure chemical ionization tandem mass spectrometry. Secondhand smoke exposure (SHS) was defined among never smokers as those with >0.05 ng/ml and <10 ng/ml of serum cotinine levels, as previously done in the NHANES cohort by the NCHS (4). SHS was also examined in former smokers who quit smoking more than 1 month ago, to ensure a complete excretion of cotinine derived from active smoking (half-life in plasma of 15-20 hours).

**Telomere length**

Telomere length was measured in whole blood samples through a quantitative polymerase chain reaction (qPCR). Samples were assayed in triplicates and telomere length was measured relative to a standard reference DNA (T/S). The T/S ratio was converted to base pairs (bp) according to bp=3,274+2,413*(T/S). More details about telomere length measurement are reported on the NHANES website (1).

**Statistical analyses**

For the statistical analyses, we filtered out participants top-coded as aged ≥85 years (n=130), those with sex mismatch between reported gender and epigenetic-predicted sex (n=56), participants without data about self-reported smoking (n=6), and never cigarette smokers who ever smoked pipe or cigar (n=20), leaving n=2,300 participants for subsequent analyses. Potential confounders and precision variables were identified *a priori* and included chronological age in years (RIDAGEYR), chronological age squared, self-reported gender (male *vs.* female; RIAGENDR), self-identified race/ethnicity (Non-Hispanic White, Mexican American, Other Hispanic, Non-Hispanic Black, Other Race/Multi-Racial; RIDRETH1), body mass index (BMXBMI), poverty-to-income ratio (INDFMPIR), education (less than high school, high school diploma or General Educational Development, greater than high school education; DMDEDUC), and survey cycles (1999-2000 and 2001-2002). Raw epigenetic age estimates, while controlling for age as a covariate, were analyzed as previously recommended for studies of epigenetic accelerated aging (5). We examined the association between smoking status, time since smoking cessation, smoking intensity, and SHS with epigenetic clocks through survey-weighted linear regression models adjusted for the described confounders. Generalized regression models were conducted using the *survey* package to account for participant sample weights and the NHANES survey design (6).

Additionally, we stratified former smokers according to time since smoking cessation (from ≤5 to >30 years) and current smokers according to smoking intensity (from ≤10 to

>25 packs in the last month) and tested for the association of epigenetic aging compared to never smokers. We also tested for the association of telomere length estimated by qPCR with smoking exposures. Multiple comparisons were adjusted using a false discovery rate (FDR)<5% considering the number of epigenetic clocks evaluated. Statistical differences between clinical or demographic variables between groups were examined using a Mann-Whitney U test. All analyses were conducted in R (v 4.4.1) (7).

Estimates from the model are represented in increases or decreases in the units of each clock, which are years for HorvathAge, HannumAge, SkinBloodAge, PhenoAge, GrimAge2, ZhangAge, LinAge, WeidnerAge, and VidalBraloAge; kilobases (kb) of methylation-predicted telomere lengths for DNAmTL; years of physiological decline occurring per 12 months of calendar time (pace of aging) for DunedinPoAm, where a score of 1 represents an average pace of aging; and average beta values in CpGs that represent a relative estimate of the number of stem cell divisions per stem cell for epiTOC. To help the interpretability of some results, years were converted to months (*12), weeks (*52), or days (*365), kb to bp (*1000), and packs of cigarettes to individual cigarettes (*20).

**Sensitivity analyses**

We conducted sensitivity analyses for all epigenetic clocks associations by further adjusting the models for blood cell counts ($CD8^+$ T cells, $CD4^+$ T cells, NK cells, B cells, monocytes, and neutrophils) to avoid any confounder effect of cell heterogeneity. Furthermore, we used cotinine levels as a measurable biomarker to validate the association of epigenetic aging with self-reported data about smoking status and intensity. Finally, to inspect whether any significant association with GrimAge2 may be driven by other mortality predictors rather than predicted pack years of smoking, we tested for the association of smoking exposures with all GrimAge2 components.

### SUPPLEMENTARY FIGURES

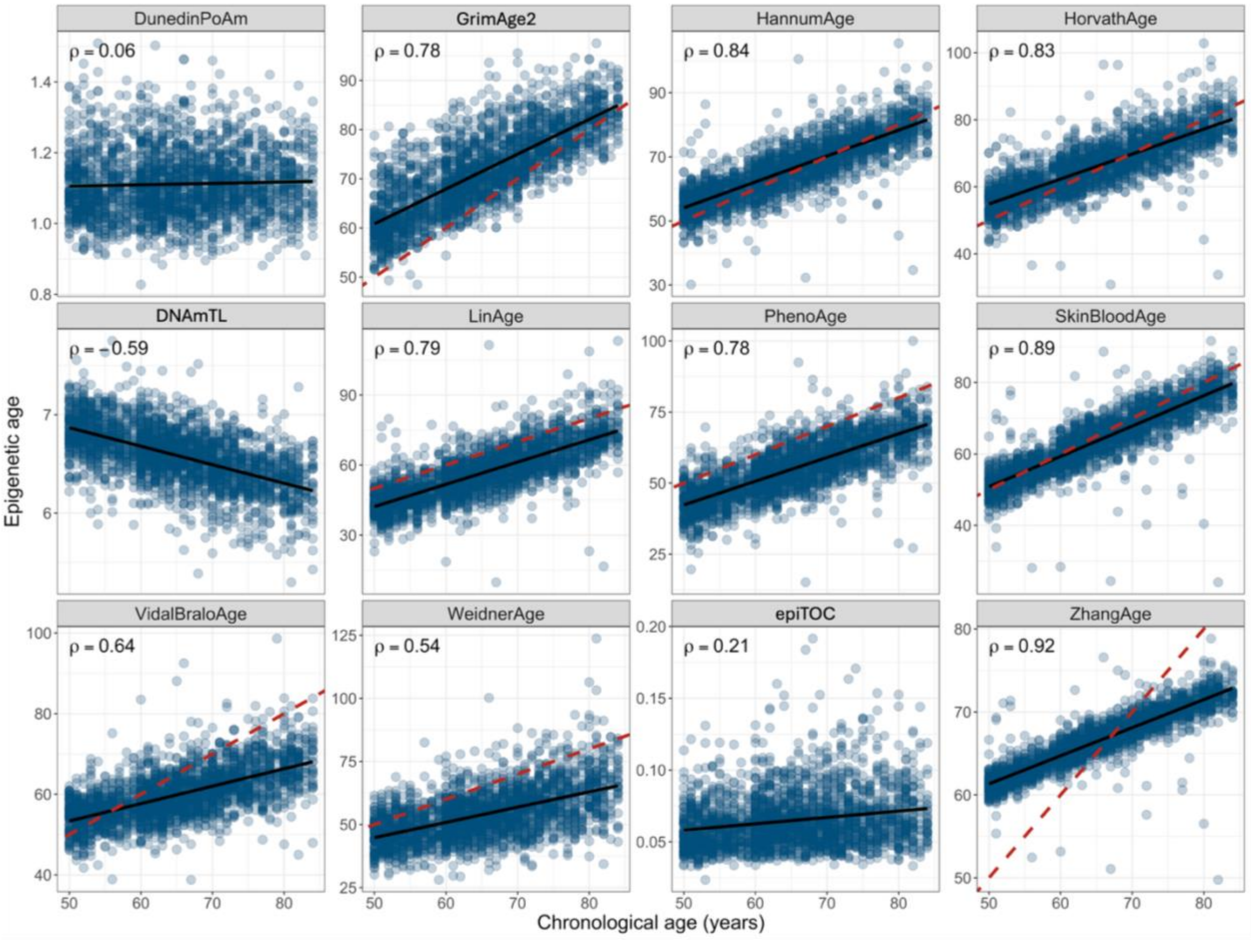


**Figure S1.** Scatter plot for the correlation between chronological age (x-axis) and epigenetic age (y-axis). Epigenetic age is measured by the HorvathAge, HannumAge, SkinBloodAge, PhenoAge, GrimAge2, ZhangAge, LinAge, WeidnerAge, and VidalBraloAge clocks in years; by the DNAmTL in kilobases; by the DunedinPoAm in unit differences of the pace of aging (a score of 1 represents an average aging pace); and by the epiTOC in average beta values in CpGs that represent a relative estimate of the number of stem cell divisions per stem cell. The Spearman correlation coefficient is represented in the upper left corner of each plot, and the black line represents the slope of the correlation. The red dashed line represents the 1:1 relationship (only for those epigenetic clocks providing estimates in years). Highest correlations were observed for chronological age clocks, such as the HorvathAge ($\rho$=0.83), HannumAge ($\rho$=0.84), SkinBloodAge ($\rho$=0.89), ZhangAge ($\rho$=0.92), and LinAge ($\rho$=0.79), and lifespan and health span clocks, such as GrimAge2 ($\rho$=0.78) and PhenoAge ($\rho$=0.78). Moderate positive correlations were detected for the VidalBraloAge ($\rho$=0.64) and WeidnerAge ($\rho$=0.54), while DNAm-predicted telomere length showed a moderate negative correlation (DNAmTL, $\rho$=-0.59). Chronological age was poorly correlated with DNAm-based predictions of mitotic cell division (epiTOC, $\rho$=0.21), and it was not correlated with the pace of aging (DunedinPoAm, $\rho$=0.06).
